# Analysis of Hypertension in Peru: Prevalence, Associated Factors, Knowledge, Management and Control, 2014-2022

**DOI:** 10.1101/2024.04.22.24306187

**Authors:** Víctor Juan Vera-Ponce, Fiorella E. Zuzunaga-Montoya, Luisa Erika Milagros Vásquez-Romero, Joan A. Loayza-Castro, Cori Raquel Iturregui Paucar, Mario J. Valladares-Garrido, Enrique Vigil-Ventura

**Affiliations:** Instituto de Investigación de Enfermedades Tropicales, Universidad Nacional Toribio Rodríguez de Mendoza de Amazonas (UNTRM), Amazonas, Perú; Facultad de Medicina (FAMED), Universidad Nacional Toribio Rodríguez de Mendoza de Amazonas (UNTRM), Amazonas, Perú; Universidad Tecnológica del Perú, Lima, Perú; Universidad Continental Lima, Perú; Oficina de Epidemiología, Hospital Regional Lambayeque, Chiclayo, Perú

**Author notes:** Correspondence, Víctor Juan Vera-Ponce, Fiorella E. Zuzunaga-Montoya,; Luisa Erika Milagros Vásquez Romero,; Joan A. Loayza-Castro,; Cori Raquel Iturregui Paucar,; Mario J. Valladares-Garrido,; Enrique Vigil-Ventura. Instituto de Enfermedades Tropicales, Universidad Nacional Toribio Rodriguez de Mendonza.

**Keywords:** Hypertension, Prehypertension, Arterial Pressure, Public Health, Awareness, Treatment (source: MeSH NLM)

## Abstract

**Introduction:** Hypertension (HTN), prehypertension, and High Blood Pressure (HBP) represent a chronic medical condition of growing global concern. Highlighting the importance of Awareness, medication, and Control of HTN is essential. A significant proportion of hypertensive patients are unaware of their condition, hindering proper and early treatment. Among those diagnosed, not all receive or follow pharmacological treatment, and an even smaller percentage achieves effective blood pressure control.

**Objective:** The main objectives include determining the prevalence, trends, and associated factors of HTN, as well as investigating prehypertension and the variability in HBP levels during the same period. Special attention will be given to self-reported, medicated, and Controlled hypertension, analyzing its evolution and associated factors.

**Methods:** We conducted an analytical cross-sectional study using the National Survey of Demographic and Family Health database between 2014 and 2022. Our study included individuals aged 18 years and older, as the definitions of HTN apply to people from this age onwards.

**Results:** The study revealed an overall HTN, prehypertension, and HBP prevalence of 20.76%, 33.44%, and 33.44%, respectively, with annual variations. An increasing trend in the prevalence of known, medicated, and controlled HTN was observed, with factors such as gender, age, educational level, and geographical region significantly influencing it.

**Conclusions:** The investigation into HTN in Peru revealed an expanding occurrence of HTN, prehypertension, and HBP, emphasizing the necessity for early identification, suitable treatment, and effective management of HTN. A significant gap is observed between the condition’s awareness and effective management, highlighting the urgent need for public health strategies that address both the prevention and treatment of HTN in Peru.

## Introduction

Hypertension (HTN) represents a chronic medical condition of growing global concern, characterized by a sustained increase in blood pressure above normal levels ^(1)^. Public health faces a formidable obstacle from this condition, especially in developing nations where its increasing prevalence presents a looming threat^(2)^. HTN, which disproportionately impacts women over men and is more prevalent in rural locales, has been associated with severe complications such as cardiovascular ailments, stroke incidents, retinopathy, and nephropathy. While noncompliance with antihypertensive regimens remains a principal factor behind insufficient blood pressure management, it consequently raises the frequency of complications and deaths attributable to the condition ^(3)^.

Globally, it is estimated that around one billion one hundred thirty million people endure hypertension, constituting roughly sixteen percent of all grown humans across the planet ^(4)^. In the United States, where approximately one-third of all adults experience this condition, the prevalence of suffering from such an ailment has become distressingly common. While estimates suggest the broad prevalence of this condition across Latin American adults ranges from 20 to 30 percent ^(5)^, Peru has explicitly seen a combined rate of roughly 22% among its population ^(6)^. These figures underscore the importance of addressing HTN as a critical public health issue in various regions.

A timely diagnosis coupled with precision in identifying HTN is paramount to forestalling grave issues and bettering one’s standard of living. In Peru, access to health services and regular health check-ups are essential for timely diagnosis. Upon receiving a diagnosis, patients must undertake suitable treatments that encompass lifestyle modifications and antihypertensive pharmaceuticals, strictly adhering to them diligently and aiming to govern the condition efficaciously ^(7,8)^.

Highlighting the importance of Awareness, medicated, and Control HTN is essential. A significant proportion of hypertensive patients are unaware of their condition, hindering proper and early treatment. Among those diagnosed, not all receive or follow pharmacological treatment, and an even smaller percentage achieve effective blood pressure control ^(5,9,10)^.

This study aims to provide a comprehensive overview of the HTN situation in Peru between 2014 and 2022. The main objectives include determining the prevalence, trends, and associated factors of HTN and investigating prehypertension and the variability in high blood pressure (HBP) levels during the same period. Special attention will be given to self-reported, medicated, and Controlled hypertension, analyzing its evolution and associated factors. By leveraging a national database to facilitate an exhaustive examination of hypertension’s prevalence, diagnosis, treatment, and management in Peru, this study hopes to furnish a nuanced understanding of its dynamics within the country to illuminate valuable perspectives for enhancing public health strategies.

## Material and Methods

### Design

We conducted an analytical cross-sectional study using a database from the National Survey of Demographic and Family Health (ENDES) between 2014 and 2022 ^(11)^. Our study followed the STROBE guidelines to ensure comprehensive reporting of observational studies in epidemiology ^(12)^.

### Population, Eligibility Criteria, and Sample

The survey was carried out nationwide and included Peruvians aged 15 to 49 from urban and rural areas across 24 departments in Peru. ENDES used a two-stage, stratified sampling method independently probabilistic for rural and urban areas ^(11)^. Our study included individuals aged 18 years and older, as the definitions of HTN apply to people from this age onwards ^(1)^. We excluded all participants who did not have their blood pressure measured.

### Assessment of Consistency and Plausibility of Measurements

To reasonably ensure that solely practical blood pressure readings were incorporated into the investigations, parameters were established mirroring the methodology of earlier related analyses, necessitating that determinations of both Systolic Blood Pressure (SBP) and Diastolic Blood Pressure (DBP) remain within set limits: measurements of SBP were required to fall between 70 mmHg and 270 mmHg, and readings of DBP between 50 mmHg and 150 mmHg. Measurements that did not meet these plausibility criteria were excluded from subsequent analyses ^(13)^.

### Variables and Measurement

The main variables were as follows:

- HTN was defined as those who had SBP ≥ 140 mmHg or DBP ≥ 90 mmHg or had a self-reported previous diagnosis of HTN ^(1)^.
- Prehypertension was defined as those individuals whose SBP was in the range of 120-139 mmHg or whose DBP was in the range of 80-89 mmHg and who had not been previously diagnosed with hypertension ^(14)^.
- HBP was considered in individuals whose SBP was 130-139 mmHg or whose DBP was 80-89 mmHg and who had not been previously diagnosed with HTN ^(1)^.
- Hypertensive patients aware of their condition were those who reported having received a prior diagnosis of hypertension.
- Participants were aware of their HTN status and self-reported that they were taking appropriate antihypertensive medication.
- Participants were aware of their HTN status, reported taking appropriate antihypertensive medication, and had average blood pressure values (SBP <140 mmHg and DBP <90 mmHg).

The study considered sex (female or male), age group (ages 20 to 35, 36 to 59, 60 to 69, and 70 or older), marital situation (being partnered or single), region (residing in Metropolitan Lima, elsewhere along the coast, in the highlands, or jungle areas), wealth index (from poorest to most affluent), an education level (having no schooling or primary education versus secondary education or higher), living in an urban or rural local, physical disability (yes or no), daily fruit and vegetable consumption above five portions (yes or no), presence of type 2 diabetes mellitus (T2DM) (yes or no), altitude ranges (0 to 499, 500 to 1499, 1500 to 2999, 3000 or more), race (Other, Quechua, Aymara, Native or Indigenous Amazonian, Negroid/Black, White/Caucasian or Mestizo), nutritional status, abdominal obesity (yes or no), Smoker Status and Alcohol consumption ^(15)^.

The race variable was evaluated by self-reporting: Because of your ancestors and customs, ¿do you consider yourself? Giving the alternatives above as options.

Abdominal obesity was evaluated according to the Cholesterol Education Program Adult Treatment Panel III (WC-ATPIII) criteria (WC ≥ 102 cm in men and abdominal waist in women ≥ 88 cm ^(16)^). The WHO classification established the cut-off points that indicate nutritional status according to BMI. The cut-off points for adults were: average weight (< 25 kg/m2), overweight (BMI between 25 and 29.9 kg/m2), and obesity (BMI ≥ 30 kg/m2), Smoker Status was obtained from the self-report of the interviewee, classified as never smoked, ex-smoker, or currently smoker ^(17)^. Alcohol consumption was also defined by self-report and was classified as Never or did not use in the last 12 months; consumption was not excessive (≥ one occasion in the last 30 days, but <5 drinks in the previous 12 months). Men or <4 drinks in women) and excessive drinking (≥1 occasion in the last 30 days and ≥ five drinks in men or ≥ four drinks in women) ^(18)^.

### Procedures

To reliably and accurately gauge systolic and diastolic blood pressure levels, a standardized process was instituted to guarantee consistent measurement outcomes. The blood pressure levels of the participants were collected using a digital sphygmomanometer (OMRON, model HEM-713), and two types of cuffs were used, adapted to the arm circumference of each individual: standard arm (220–320 mm) and giant arm (330– 430 mm).

The measurements were carefully conducted in a regulated setting that guaranteed the participants to be at ease, seated with their right arm laying flat on a level surface at the height of their heart. The initial measurement occurred after a 5-minute interval of relaxation, while the secondary determination took place two minutes following the former; this timeframe between the two estimations enabled cardiovascular parameters to achieve equilibrium. Subsequently, the mean of both the systolic and diastolic blood pressure readings for each individual was derived, and this mean numerical value served as the basis for the analyses above. This procedure furnishes clinicians with a more reliable means of assessment by stabilizing blood pressure fluctuations and capturing a more precise portrait of an individual’s typical blood pressure levels.

In addition to gathering self-reported data on whether participants had ever received a prior diagnosis of hypertension or were currently taking medication to treat high blood pressure, the National Survey of Demographic and Family Health also collected various pertinent sociodemographic information needed to inform the research.

### Statistical Analysis

The R software version 4.03 will perform various statistical analyses in the present study. Firstly, a descriptive analysis of all variables will be conducted, providing an overview of the dataset’s characteristics.

Subsequently, a bivariate analysis will be performed for HTN, Prehypertension, and HBP. Additionally, a specific bivariate analysis will be conducted for Awareness HTN, Treatment HTN, and Control HTN, exploring the relationships between these categories and other study variables.

A Poisson regression model with robust variance was implemented to delve deeper into the analysis of associated factors. This model will allow the calculation of Prevalence Ratios (PR) and analyze the associated factors of HTN, Prehypertension, and HBP. Similarly, a regression analysis will evaluate awareness of HTN, treatment of HTN, control of HTN, and associated factors.

Regarding the visualization of the results, various trend graphs covering 2014 to 2022 will be generated. Graphs for HTN, Prehypertension, and HBP will be developed globally and stratified by sex. Additionally, graphs will be created to visualize the trend of Awareness HTN, Treatment HTN, and Control HTN.

### Ethical Considerations

In this study, data that are freely and publicly accessible and anonymized, meaning they do not contain information that allows the personal identification of participants, thus eliminating any ethical risk to them, have been used. The National Institute of Statistics and Informatics collects ENDES’s annual data. This Peruvian government entity ensures informed consent is obtained from participants to collect the necessary information through the survey. For adult participants aged 18 and over, consent is directly collected. In the case of participants under 18, consent is requested from one of their parents or the responsible adult, thus ensuring ethics in evaluating minors.

## Results

### Participants

We included a total of 275,759 participants. The gender distribution was balanced, with 52.28% being women. 17.53% were older adults. Regarding educational level, 33.48% had higher education. On the other hand, 23.31% resided in rural areas. As for harmful habits, 10.94% of participants reported current smoking, while 2.70% reported excessive alcohol consumption. Regarding fruit and vegetable intake, only 8.98% consumed five or more daily servings. The total descriptive characteristics can be visualized in Table 1.

**Table 1.**
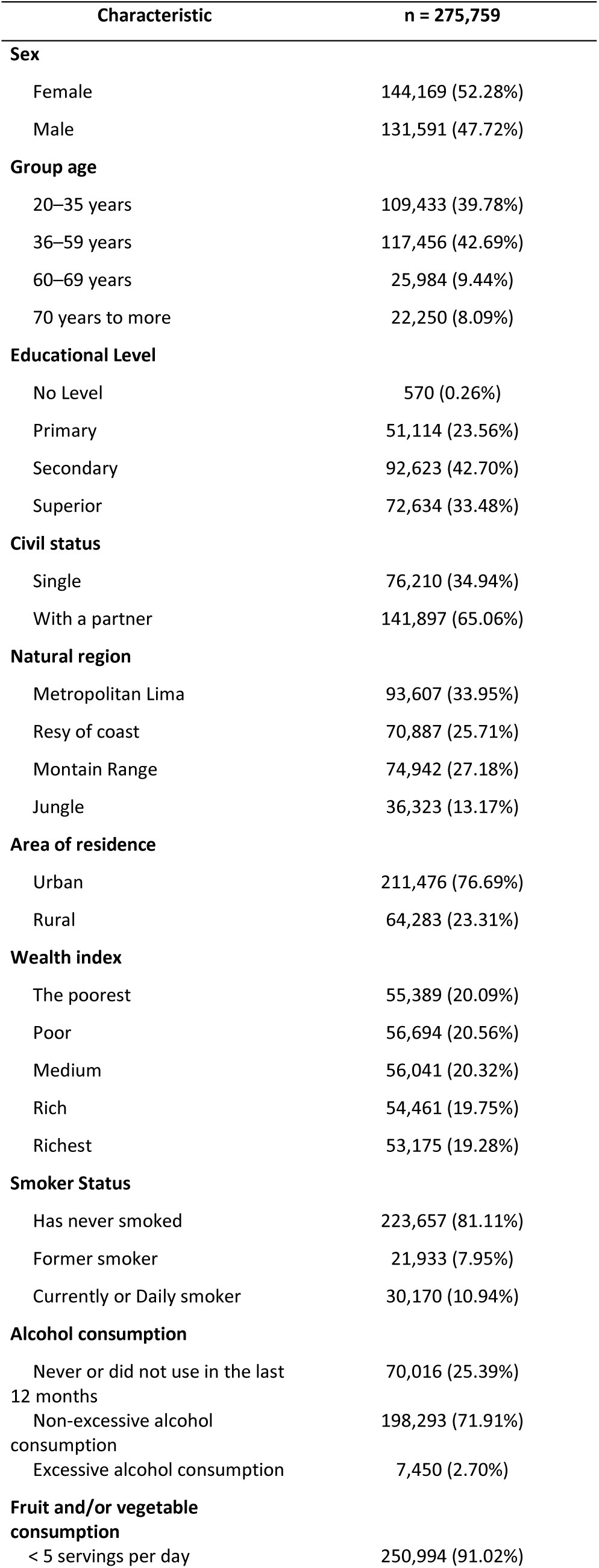
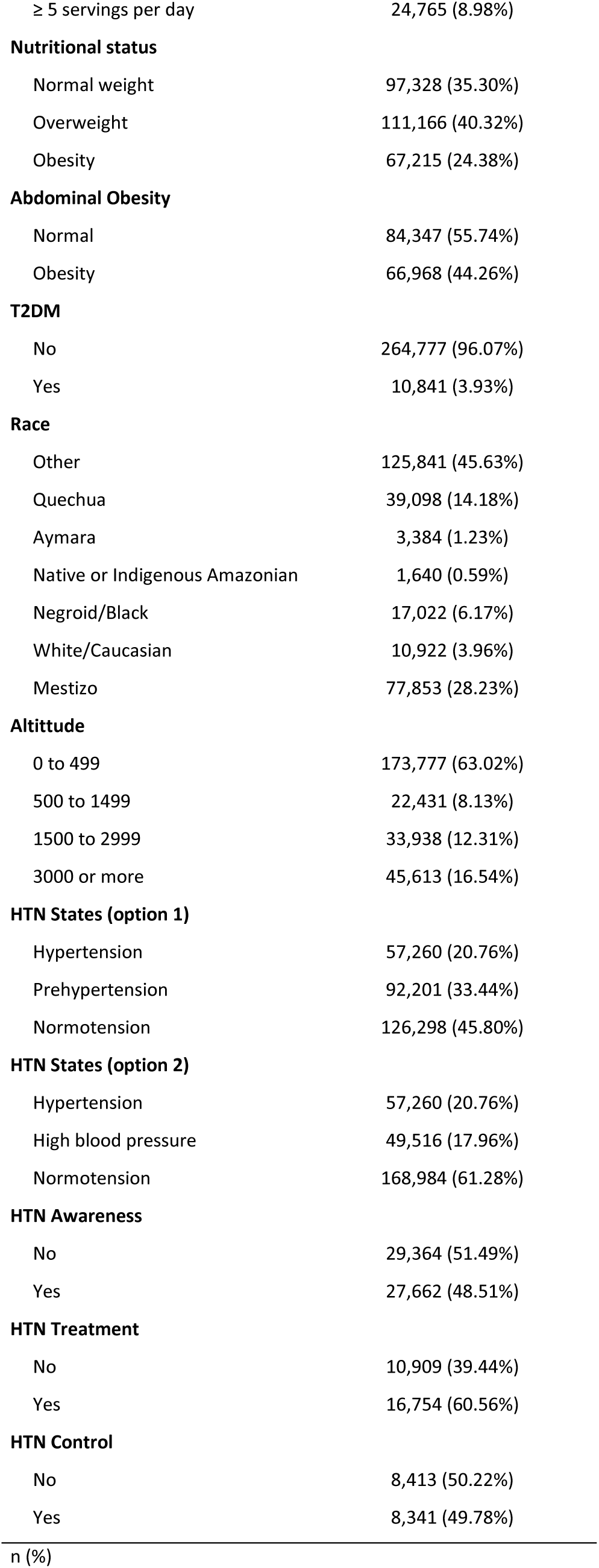
Descriptive characteristics of the study sample.

| Characteristic | n = 275,759 |
| --- | --- |
| **Sex** |  |
| Female | 144,169 (52.28%) |
| Male | 131,591 (47.72%) |
| **Group age** |  |
| 20–35 years | 109,433 (39.78%) |
| 36–59 years | 117,456 (42.69%) |
| 60–69 years | 25,984 (9.44%) |
| 70 years to more | 22,250 (8.09%) |
| **Educational Level** |  |
| No Level | 570 (0.26%) |
| Primary | 51,114 (23.56%) |
| Secondary | 92,623 (42.70%) |
| Superior | 72,634 (33.48%) |
| **Civil status** |  |
| Single | 76,210 (34.94%) |
| With a partner | 141,897 (65.06%) |
| **Natural region** |  |
| Metropolitan Lima | 93,607 (33.95%) |
| Resy of coast | 70,887 (25.71%) |
| Montain Range | 74,942 (27.18%) |
| Jungle | 36,323 (13.17%) |
| **Area of residence** |  |
| Urban | 211,476 (76.69%) |
| Rural | 64,283 (23.31%) |
| **Wealth index** |  |
| The poorest | 55,389 (20.09%) |
| Poor | 56,694 (20.56%) |
| Medium | 56,041 (20.32%) |
| Rich | 54,461 (19.75%) |
| Richest | 53,175 (19.28%) |
| **Smoker Status** |  |
| Has never smoked | 223,657 (81.11%) |
| Former smoker | 21,933 (7.95%) |
| Currently or Daily smoker | 30,170 (10.94%) |
| **Alcohol consumption** |  |
| Never or did not use in the last 12 months | 70,016 (25.39%) |
| Non-excessive alcohol consumption | 198,293 (71.91%) |
| Excessive alcohol consumption | 7,450 (2.70%) |
| **Fruit and/or vegetable consumption** |  |
| < 5 servings per day | 250,994 (91.02%) |
| ≥ 5 servings per day | 24,765 (8.98%) |
| **Nutritional status** |  |
| Normal weight | 97,328 (35.30%) |
| Overweight | 111,166 (40.32%) |
| Obesity | 67,215 (24.38%) |
| **Abdominal Obesity** |  |
| Normal | 84,347 (55.74%) |
| Obesity | 66,968 (44.26%) |
| **T2DM** |  |
| No | 264,777 (96.07%) |
| Yes | 10,841 (3.93%) |
| **Race** |  |
| Other | 125,841 (45.63%) |
| Quechua | 39,098 (14.18%) |
| Aymara | 3,384 (1.23%) |
| Native or Indigenous Amazonian | 1,640 (0.59%) |
| Negroid/Black | 17,022 (6.17%) |
| White/Caucasian | 10,922 (3.96%) |
| Mestizo | 77,853 (28.23%) |
| **Altitude** |  |
| 0 to 499 | 173,777 (63.02%) |
| 500 to 1499 | 22,431 (8.13%) |
| 1500 to 2999 | 33,938 (12.31%) |
| 3000 or more | 45,613 (16.54%) |
| **HTN States (option 1)** |  |
| Hypertension | 57,260 (20.76%) |
| Prehypertension | 92,201 (33.44%) |
| Normotension | 126,298 (45.80%) |
| **HTN States (option 2)** |  |
| Hypertension | 57,260 (20.76%) |
| High blood pressure | 49,516 (17.96%) |
| Normotension | 168,984 (61.28%) |
| **HTN Awareness** |  |
| No | 29,364 (51.49%) |
| Yes | 27,662 (48.51%) |
| **HTN Treatment** |  |
| No | 10,909 (39.44%) |
| Yes | 16,754 (60.56%) |
| **HTN Control** |  |
| No | 8,413 (50.22%) |
| Yes | 8,341 (49.78%) |

**Table 2.**
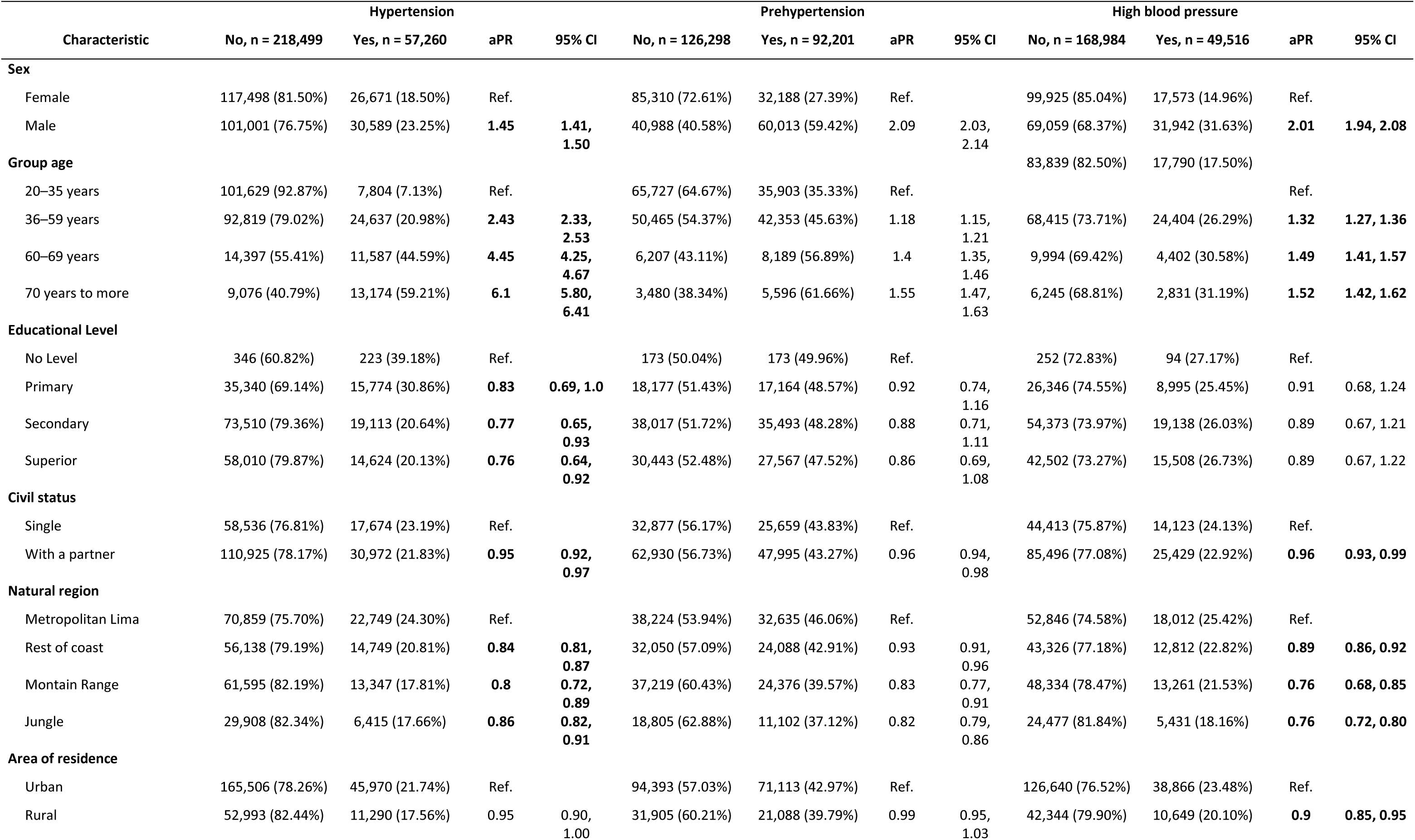
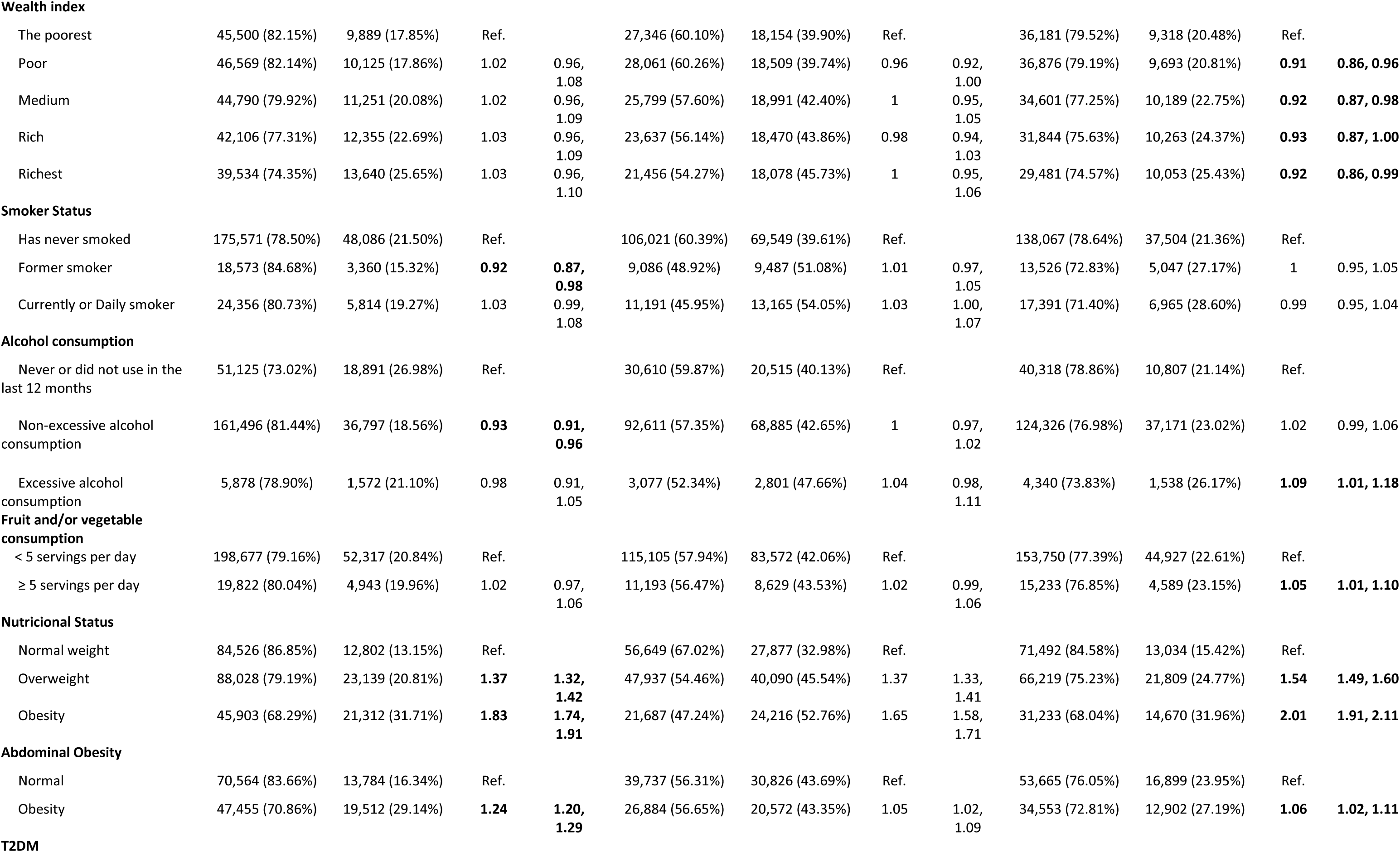
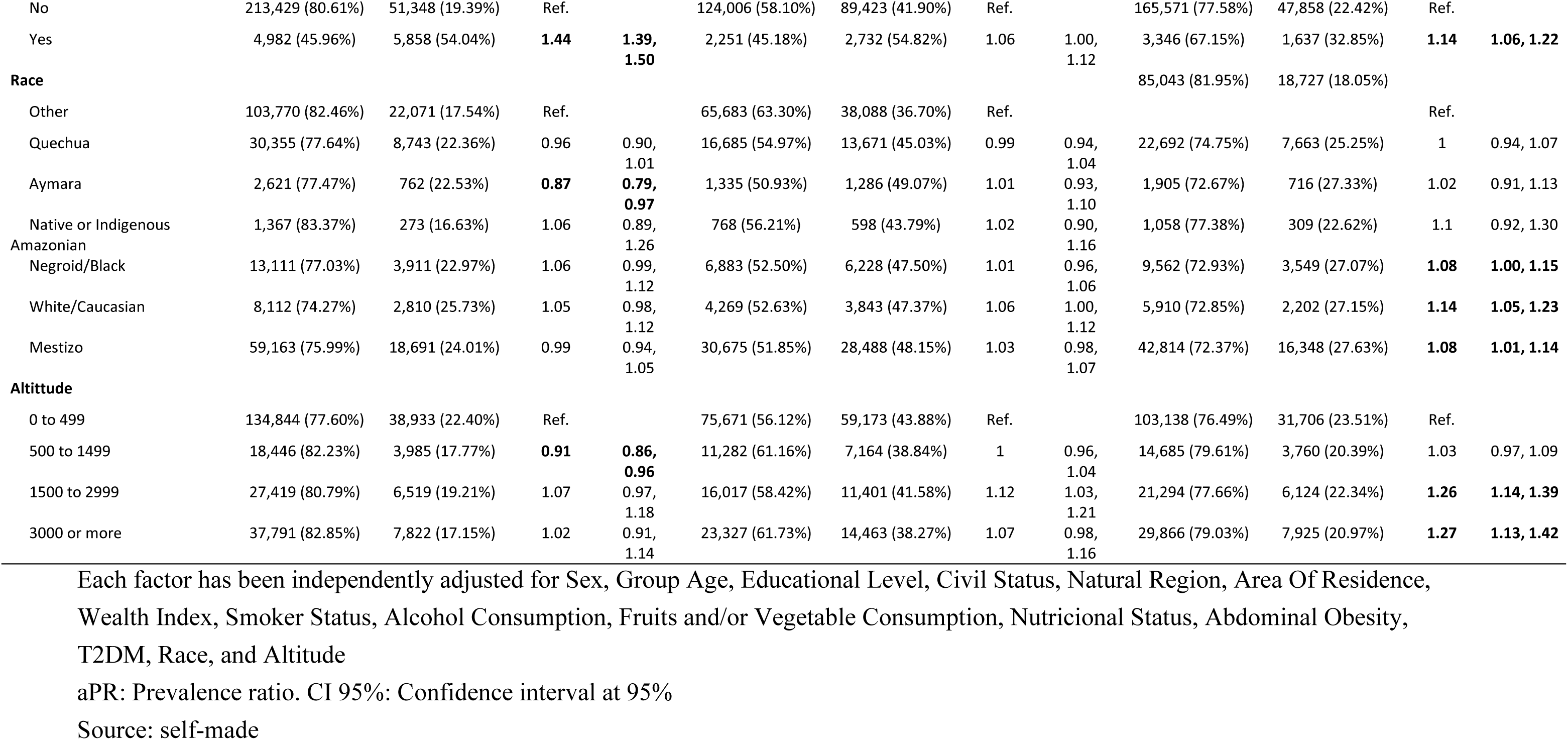
Bivariate analysis and factors associated with HTN, prehypertension and HBP.

| Characteristic | Hypertension |  |  |  | Prehypertension |  |  |  | High blood pressure |  |  |  |
| --- | --- | --- | --- | --- | --- | --- | --- | --- | --- | --- | --- | --- |
|  | No, n = 218,499 | Yes, n = 57,260 | aPR | 95% CI | No, n = 126,298 | Yes, n = 92,201 | aPR | 95% CI | No, n = 168,984 | Yes, n = 49,516 | aPR | 95% CI |
| **Sex** |  |  |  |  |  |  |  |  |  |  |  |  |
| Female | 117,498 (81.50%) | 26,671 (18.50%) | Ref. |  | 85,310 (72.61%) | 32,188 (27.39%) | Ref. |  | 99,925 (85.04%) | 17,573 (14.96%) | Ref. |  |
| Male | 101,001 (76.75%) | 30,589 (23.25%) | **1.45** | **1.41, 1.50** | 40,988 (40.58%) | 60,013 (59.42%) | 2.09 | 2.03, 2.14 | 69,059 (68.37%) | 31,942 (31.63%) | **2.01** | **1.94, 2.08** |
| **Group age** |  |  |  |  |  |  |  |  | 83,839 (82.50%) | 17,790 (17.50%) |  |  |
| 20–35 years | 101,629 (92.87%) | 7,804 (7.13%) | Ref. |  | 65,727 (64.67%) | 35,903 (35.33%) | Ref. |  |  |  | Ref. |  |
| 36–59 years | 92,819 (79.02%) | 24,637 (20.98%) | **2.43** | **2.33, 2.53** | 50,465 (54.37%) | 42,353 (45.63%) | 1.18 | 1.15, 1.21 | 68,415 (73.71%) | 24,404 (26.29%) | **1.32** | **1.27, 1.36** |
| 60–69 years | 14,397 (55.41%) | 11,587 (44.59%) | **4.45** | **4.25, 4.67** | 6,207 (43.11%) | 8,189 (56.89%) | 1.4 | 1.35, 1.46 | 9,994 (69.42%) | 4,402 (30.58%) | **1.49** | **1.41, 1.57** |
| 70 years to more | 9,076 (40.79%) | 13,174 (59.21%) | **6.1** | **5.80, 6.41** | 3,480 (38.34%) | 5,596 (61.66%) | 1.55 | 1.47, 1.63 | 6,245 (68.81%) | 2,831 (31.19%) | **1.52** | **1.42, 1.62** |
| **Educational Level** |  |  |  |  |  |  |  |  |  |  |  |  |
| No Level | 346 (60.82%) | 223 (39.18%) | Ref. |  | 173 (50.04%) | 173 (49.96%) | Ref. |  | 252 (72.83%) | 94 (27.17%) | Ref. |  |
| Primary | 35,340 (69.14%) | 15,774 (30.86%) | **0.83** | **0.69, 1.0** | 18,177 (51.43%) | 17,164 (48.57%) | 0.92 | 0.74, 1.16 | 26,346 (74.55%) | 8,995 (25.45%) | 0.91 | 0.68, 1.24 |
| Secondary | 73,510 (79.36%) | 19,113 (20.64%) | **0.77** | **0.65, 0.93** | 38,017 (51.72%) | 35,493 (48.28%) | 0.88 | 0.71, 1.11 | 54,373 (73.97%) | 19,138 (26.03%) | 0.89 | 0.67, 1.21 |
| Superior | 58,010 (79.87%) | 14,624 (20.13%) | **0.76** | **0.64, 0.92** | 30,443 (52.48%) | 27,567 (47.52%) | 0.86 | 0.69, 1.08 | 42,502 (73.27%) | 15,508 (26.73%) | 0.89 | 0.67, 1.22 |
| **Civil status** |  |  |  |  |  |  |  |  |  |  |  |  |
| Single | 58,536 (76.81%) | 17,674 (23.19%) | Ref. |  | 32,877 (56.17%) | 25,659 (43.83%) | Ref. |  | 44,413 (75.87%) | 14,123 (24.13%) | Ref. |  |
| With a partner | 110,925 (78.17%) | 30,972 (21.83%) | **0.95** | **0.92, 0.97** | 62,930 (56.73%) | 47,995 (43.27%) | 0.96 | 0.94, 0.98 | 85,496 (77.08%) | 25,429 (22.92%) | **0.96** | **0.93, 0.99** |
| **Natural region** |  |  |  |  |  |  |  |  |  |  |  |  |
| Metropolitan Lima | 70,859 (75.70%) | 22,749 (24.30%) | Ref. |  | 38,224 (53.94%) | 32,635 (46.06%) | Ref. |  | 52,846 (74.58%) | 18,012 (25.42%) | Ref. |  |
| Rest of coast | 56,138 (79.19%) | 14,749 (20.81%) | **0.84** | **0.81, 0.87** | 32,050 (57.09%) | 24,088 (42.91%) | 0.93 | 0.91, 0.96 | 43,326 (77.18%) | 12,812 (22.82%) | **0.89** | **0.86, 0.92** |
| Mountain Range | 61,595 (82.19%) | 13,347 (17.81%) | **0.8** | **0.72, 0.89** | 37,219 (60.43%) | 24,376 (39.57%) | 0.83 | 0.77, 0.91 | 48,334 (78.47%) | 13,261 (21.53%) | **0.76** | **0.68, 0.85** |
| Jungle | 29,908 (82.34%) | 6,415 (17.66%) | **0.86** | **0.82, 0.91** | 18,805 (62.88%) | 11,102 (37.12%) | 0.82 | 0.79, 0.86 | 24,477 (81.84%) | 5,431 (18.16%) | **0.76** | **0.72, 0.80** |
| **Area of residence** |  |  |  |  |  |  |  |  |  |  |  |  |
| Urban | 165,506 (78.26%) | 45,970 (21.74%) | Ref. |  | 94,393 (57.03%) | 71,113 (42.97%) | Ref. |  | 126,640 (76.52%) | 38,866 (23.48%) | Ref. |  |
| Rural | 52,993 (82.44%) | 11,290 (17.56%) | 0.95 | 0.90, 1.00 | 31,905 (60.21%) | 21,088 (39.79%) | 0.99 | 0.95, 1.03 | 42,344 (79.90%) | 10,649 (20.10%) | **0.9** | **0.85, 0.95** |
| **Wealth index** |  |  |  |  |  |  |  |  |  |  |  |  |
| The poorest | 45,500 (82.15%) | 9,889 (17.85%) | Ref. |  | 27,346 (60.10%) | 18,154 (39.90%) | Ref. |  | 36,181 (79.52%) | 9,318 (20.48%) | Ref. |  |
| Poor | 46,569 (82.14%) | 10,125 (17.86%) | 1.02 | 0.96,<br>1.08 | 28,061 (60.26%) | 18,509 (39.74%) | 0.96 | 0.92,<br>1.00 | 36,876 (79.19%) | 9,693 (20.81%) | **0.91** | **0.86, 0.96** |
| Medium | 44,790 (79.92%) | 11,251 (20.08%) | 1.02 | 0.96,<br>1.09 | 25,799 (57.60%) | 18,991 (42.40%) | 1 | 0.95,<br>1.05 | 34,601 (77.25%) | 10,189 (22.75%) | **0.92** | **0.87, 0.98** |
| Rich | 42,106 (77.31%) | 12,355 (22.69%) | 1.03 | 0.96,<br>1.09 | 23,637 (56.14%) | 18,470 (43.86%) | 0.98 | 0.94,<br>1.03 | 31,844 (75.63%) | 10,263 (24.37%) | **0.93** | **0.87, 1.00** |
| Richest | 39,534 (74.35%) | 13,640 (25.65%) | 1.03 | 0.96,<br>1.10 | 21,456 (54.27%) | 18,078 (45.73%) | 1 | 0.95,<br>1.06 | 29,481 (74.57%) | 10,053 (25.43%) | **0.92** | **0.86, 0.99** |
| **Smoker Status** |  |  |  |  |  |  |  |  |  |  |  |  |
| Has never smoked | 175,571 (78.50%) | 48,086 (21.50%) | Ref. |  | 106,021 (60.39%) | 69,549 (39.61%) | Ref. |  | 138,067 (78.64%) | 37,504 (21.36%) | Ref. |  |
| Former smoker | 18,573 (84.68%) | 3,360 (15.32%) | **0.92** | **0.87,**<br>**0.98** | 9,086 (48.92%) | 9,487 (51.08%) | 1.01 | 0.97,<br>1.05 | 13,526 (72.83%) | 5,047 (27.17%) | 1 | 0.95, 1.05 |
| Currently or Daily smoker | 24,356 (80.73%) | 5,814 (19.27%) | 1.03 | 0.99,<br>1.08 | 11,191 (45.95%) | 13,165 (54.05%) | 1.03 | 1.00,<br>1.07 | 17,391 (71.40%) | 6,965 (28.60%) | 0.99 | 0.95, 1.04 |
| **Alcohol consumption** |  |  |  |  |  |  |  |  |  |  |  |  |
| Never or did not use in the last 12 months | 51,125 (73.02%) | 18,891 (26.98%) | Ref. |  | 30,610 (59.87%) | 20,515 (40.13%) | Ref. |  | 40,318 (78.86%) | 10,807 (21.14%) | Ref. |  |
| Non-excessive alcohol consumption | 161,496 (81.44%) | 36,797 (18.56%) | **0.93** | **0.91,**<br>**0.96** | 92,611 (57.35%) | 68,885 (42.65%) | 1 | 0.97,<br>1.02 | 124,326 (76.98%) | 37,171 (23.02%) | 1.02 | 0.99, 1.06 |
| Excessive alcohol consumption | 5,878 (78.90%) | 1,572 (21.10%) | 0.98 | 0.91,<br>1.05 | 3,077 (52.34%) | 2,801 (47.66%) | 1.04 | 0.98,<br>1.11 | 4,340 (73.83%) | 1,538 (26.17%) | **1.09** | **1.01, 1.18** |
| **Fruit and/or vegetable consumption** |  |  |  |  |  |  |  |  |  |  |  |  |
| < 5 servings per day | 198,677 (79.16%) | 52,317 (20.84%) | Ref. |  | 115,105 (57.94%) | 83,572 (42.06%) | Ref. |  | 153,750 (77.39%) | 44,927 (22.61%) | Ref. |  |
| ≥ 5 servings per day | 19,822 (80.04%) | 4,943 (19.96%) | 1.02 | 0.97,<br>1.06 | 11,193 (56.47%) | 8,629 (43.53%) | 1.02 | 0.99,<br>1.06 | 15,233 (76.85%) | 4,589 (23.15%) | **1.05** | **1.01, 1.10** |
| **Nutritional Status** |  |  |  |  |  |  |  |  |  |  |  |  |
| Normal weight | 84,526 (86.85%) | 12,802 (13.15%) | Ref. |  | 56,649 (67.02%) | 27,877 (32.98%) | Ref. |  | 71,492 (84.58%) | 13,034 (15.42%) | Ref. |  |
| Overweight | 88,028 (79.19%) | 23,139 (20.81%) | **1.37** | **1.32,**<br>**1.42** | 47,937 (54.46%) | 40,090 (45.54%) | 1.37 | 1.33,<br>1.41 | 66,219 (75.23%) | 21,809 (24.77%) | **1.54** | **1.49, 1.60** |
| Obesity | 45,903 (68.29%) | 21,312 (31.71%) | **1.83** | **1.74,**<br>**1.91** | 21,687 (47.24%) | 24,216 (52.76%) | 1.65 | 1.58,<br>1.71 | 31,233 (68.04%) | 14,670 (31.96%) | **2.01** | **1.91, 2.11** |
| **Abdominal Obesity** |  |  |  |  |  |  |  |  |  |  |  |  |
| Normal | 70,564 (83.66%) | 13,784 (16.34%) | Ref. |  | 39,737 (56.31%) | 30,826 (43.69%) | Ref. |  | 53,665 (76.05%) | 16,899 (23.95%) | Ref. |  |
| Obesity | 47,455 (70.86%) | 19,512 (29.14%) | **1.24** | **1.20,**<br>**1.29** | 26,884 (56.65%) | 20,572 (43.35%) | 1.05 | 1.02,<br>1.09 | 34,553 (72.81%) | 12,902 (27.19%) | **1.06** | **1.02, 1.11** |
| **T2DM** |  |  |  |  |  |  |  |  |  |  |  |  |
| No | 213,429 (80.61%) | 51,348 (19.39%) | Ref. |  | 124,006 (58.10%) | 89,423 (41.90%) | Ref. |  | 165,571 (77.58%) | 47,858 (22.42%) | Ref. |  |
| Yes | 4,982 (45.96%) | 5,858 (54.04%) | **1.44** | **1.39, 1.50** | 2,251 (45.18%) | 2,732 (54.82%) | 1.06 | 1.00, 1.12 | 3,346 (67.15%) | 1,637 (32.85%) | **1.14** | **1.06, 1.22** |
| **Race** |  |  |  |  |  |  |  |  | 85,043 (81.95%) | 18,727 (18.05%) |  |  |
| Other | 103,770 (82.46%) | 22,071 (17.54%) | Ref. |  | 65,683 (63.30%) | 38,088 (36.70%) | Ref. |  |  |  | Ref. |  |
| Quechua | 30,355 (77.64%) | 8,743 (22.36%) | 0.96 | 0.90, 1.01 | 16,685 (54.97%) | 13,671 (45.03%) | 0.99 | 0.94, 1.04 | 22,692 (74.75%) | 7,663 (25.25%) | 1 | 0.94, 1.07 |
| Aymara | 2,621 (77.47%) | 762 (22.53%) | **0.87** | **0.79, 0.97** | 1,335 (50.93%) | 1,286 (49.07%) | 1.01 | 0.93, 1.10 | 1,905 (72.67%) | 716 (27.33%) | 1.02 | 0.91, 1.13 |
| Native or Indigenous Amazonian | 1,367 (83.37%) | 273 (16.63%) | 1.06 | 0.89, 1.26 | 768 (56.21%) | 598 (43.79%) | 1.02 | 0.90, 1.16 | 1,058 (77.38%) | 309 (22.62%) | 1.1 | 0.92, 1.30 |
| Negroid/Black | 13,111 (77.03%) | 3,911 (22.97%) | 1.06 | 0.99, 1.12 | 6,883 (52.50%) | 6,228 (47.50%) | 1.01 | 0.96, 1.06 | 9,562 (72.93%) | 3,549 (27.07%) | **1.08** | **1.00, 1.15** |
| White/Caucasian | 8,112 (74.27%) | 2,810 (25.73%) | 1.05 | 0.98, 1.12 | 4,269 (52.63%) | 3,843 (47.37%) | 1.06 | 1.00, 1.12 | 5,910 (72.85%) | 2,202 (27.15%) | **1.14** | **1.05, 1.23** |
| Mestizo | 59,163 (75.99%) | 18,691 (24.01%) | 0.99 | 0.94, 1.05 | 30,675 (51.85%) | 28,488 (48.15%) | 1.03 | 0.98, 1.07 | 42,814 (72.37%) | 16,348 (27.63%) | **1.08** | **1.01, 1.14** |
| **Altitude** |  |  |  |  |  |  |  |  |  |  |  |  |
| 0 to 499 | 134,844 (77.60%) | 38,933 (22.40%) | Ref. |  | 75,671 (56.12%) | 59,173 (43.88%) | Ref. |  | 103,138 (76.49%) | 31,706 (23.51%) | Ref. |  |
| 500 to 1499 | 18,446 (82.23%) | 3,985 (17.77%) | **0.91** | **0.86, 0.96** | 11,282 (61.16%) | 7,164 (38.84%) | 1 | 0.96, 1.04 | 14,685 (79.61%) | 3,760 (20.39%) | 1.03 | 0.97, 1.09 |
| 1500 to 2999 | 27,419 (80.79%) | 6,519 (19.21%) | 1.07 | 0.97, 1.18 | 16,017 (58.42%) | 11,401 (41.58%) | 1.12 | 1.03, 1.21 | 21,294 (77.66%) | 6,124 (22.34%) | **1.26** | **1.14, 1.39** |
| 3000 or more | 37,791 (82.85%) | 7,822 (17.15%) | 1.02 | 0.91, 1.14 | 23,327 (61.73%) | 14,463 (38.27%) | 1.07 | 0.98, 1.16 | 29,866 (79.03%) | 7,925 (20.97%) | **1.27** | **1.13, 1.42** |

Each factor has been independently adjusted for Sex, Group Age, Educational Level, Civil Status, Natural Region, Area Of Residence, Wealth Index, Smoker Status, Alcohol Consumption, Fruits and/or Vegetable Consumption, Nutricional Status, Abdominal Obesity, T2DM, Race, and Altitude
aPR: Prevalence ratio. CI 95%: Confidence interval at 95%
Source: self-made

**Table 3.** Bivariate analysis and factors associated with HTN Awareness, Treatment, and Control.

| Characteristic | Awareness |  |  |  | Treatment |  |  |  | Control |  |  |  |
| --- | --- | --- | --- | --- | --- | --- | --- | --- | --- | --- | --- | --- |
|  | No, N = 29,364 | Yes, N = 27,662 | aPR | 95% CI | No, N = 10,909 | Yes, N = 16,754 | aPR | 95% CI | No, N = 8,413 | Yes, N = 8,341 | aPR | 95% CI |
| **Sex** |  |  |  |  |  |  |  |  |  |  |  |  |
| Female | 9,572 (36.05%) | 16,979 (63.95%) | Ref. |  | 6,617 (38.97%) | 10,361 (61.03%) | Ref. |  | 4,787 (46.20%) | 5,574 (53.80%) | Ref. |  |
| Male | 19,793 (64.94%) | 10,684 (35.06%) | **0.64** | **0.61, 0.67** | 4,291 (40.17%) | 6,392 (59.83%) | **1.02** | **0.96, 1.07** | 3,626 (56.72%) | 2,766 (43.28%) | **0.74** | **0.69, 0.80** |
| **Group age** |  |  |  |  |  |  |  |  |  |  |  |  |
| 20–35 years | 5,451 (70.33%) | 2,300 (29.67%) | Ref. |  | 1,709 (74.32%) | 591 (25.68%) | Ref. |  | 104 (17.67%) | 486 (82.33%) | Ref. |  |
| 36–59 years | 13,789 (56.17%) | 10,759 (43.83%) | **1.48** | **1.36, 1.61** | 5,397 (50.16%) | 5,363 (49.84%) | **2.05** | **1.75, 2.41** | 2,247 (41.90%) | 3,116 (58.10%) | **0.78** | **0.65, 0.96** |
| 60–69 years | 5,117 (44.26%) | 6,444 (55.74%) | **1.89** | **1.74, 2.06** | 2,037 (31.61%) | 4,407 (68.39%) | **2.66** | **2.27, 3.14** | 2,251 (51.07%) | 2,156 (48.93%) | **0.7** | **0.58, 0.86** |
| 70 years to more | 4,986 (38.04%) | 8,122 (61.96%) | **2.29** | **2.10, 2.50** | 1,744 (21.47%) | 6,378 (78.53%) | **3.07** | **2.62, 3.63** | 3,801 (59.60%) | 2,577 (40.40%) | **0.63** | **0.52, 0.78** |
| **Educational Level** |  |  |  |  |  |  |  |  |  |  |  |  |
| No Level | 111 (50.27%) | 110 (49.73%) | Ref. |  | 47 (42.76%) | 63 (57.24%) | Ref. |  | 39 (62.92%) | 23 (37.08%) | Ref. |  |
| Primary | 7,242 (46.10%) | 8,468 (53.90%) | 1.16 | 0.90, 1.54 | 3,119 (36.84%) | 5,348 (63.16%) | 0.91 | 0.66, 1.31 | 3,009 (56.25%) | 2,340 (43.75%) | 1.14 | 0.69, 2.07 |
| Secondary | 10,978 (57.62%) | 8,074 (42.38%) | 1.21 | 0.93, 1.60 | 3,413 (42.28%) | 4,660 (57.72%) | 0.92 | 0.66, 1.32 | 2,300 (49.35%) | 2,360 (50.65%) | 1.27 | 0.77, 2.32 |
| Superior | 7,934 (54.40%) | 6,651 (45.60%) | 1.29 | 1.0, 1.72 | 2,229 (33.52%) | 4,422 (66.48%) | 1.05 | 0.76, 1.51 | 2,074 (46.90%) | 2,348 (53.10%) | 1.41 | 0.85, 2.57 |
| **Civil status** |  |  |  |  |  |  |  |  |  |  |  |  |
| Single | 9,081 (51.59%) | 8,522 (48.41%) | Ref. |  | 3,178 (37.29%) | 5,344 (62.71%) | — | — | 2,758 (51.60%) | 2,586 (48.40%) | — | — |
| With a partner | 15,553 (50.40%) | 15,306 (49.60%) | 1.01 | 0.97, 1.05 | 5,893 (38.50%) | 9,413 (61.50%) | **0.95** | **0.91, 0.99** | 4,760 (50.57%) | 4,653 (49.43%) | 1.01 | 0.95, 1.08 |
| **Natural region** |  |  |  |  |  |  |  |  |  |  |  |  |
| Metropolitan Lima | 12,481 (55.06%) | 10,189 (44.94%) | Ref. |  | 3,333 (32.71%) | 6,856 (67.29%) | Ref. |  | 3,698 (53.94%) | 3,158 (46.06%) | Ref. |  |
| Rest of coast | 7,307 (49.69%) | 7,397 (50.31%) | **1.12** | **1.07, 1.18** | 2,519 (34.05%) | 4,878 (65.95%) | 1.05 | 0.99, 1.12 | 2,455 (50.33%) | 2,423 (49.67%) | **1.12** | **1.03, 1.22** |
| Mountain Range | 6,733 (50.76%) | 6,531 (49.24%) | **1.42** | **1.22, 1.67** | 3,545 (54.28%) | 2,986 (45.72%) | 1.13 | 0.91, 1.41 | 1,354 (45.34%) | 1,632 (54.66%) | 1.17 | 0.85, 1.62 |
| Jungle | 2,844 (44.51%) | 3,546 (55.49%) | **1.43** | **1.33, 1.54** | 1,512 (42.64%) | 2,034 (57.36%) | 1.09 | 1.00, 1.20 | 907 (44.57%) | 1,127 (55.43%) | **1.17** | **1.03, 1.33** |
| **Area of residence** |  |  |  |  |  |  |  |  |  |  |  |  |
| Urban | 23,515 (51.32%) | 22,304 (48.68%) | Ref. |  | 8,219 (36.85%) | 14,085 (63.15%) | Ref. |  | 7,207 (51.17%) | 6,878 (48.83%) | Ref. |  |
| Rural | 5,849 (52.19%) | 5,358 (47.81%) | 1.06 | 0.98, 1.15 | 2,690 (50.19%) | 2,669 (49.81%) | 1.1 | 0.99, 1.22 | 1,206 (45.19%) | 1,463 (54.81%) | 1.09 | 0.94, 1.27 |
| **Wealth index** |  |  |  |  |  |  |  |  |  |  |  |  |
| The poorest | 5,381 (54.83%) | 4,434 (45.17%) | Ref. |  | 2,325 (52.44%) | 2,109 (47.56%) | Ref. |  | 992 (47.06%) | 1,116 (52.94%) | Ref. |  |
| Poor | 5,360 (53.22%) | 4,712 (46.78%) | 1.01 | 0.93, 1.10 | 2,228 (47.27%) | 2,485 (52.73%) | 1.17 | 1.04, 1.31 | 1,203 (48.41%) | 1,282 (51.59%) | 0.98 | 0.83, 1.15 |
| Medium | 5,821 (51.93%) | 5,388 (48.07%) | 1.04 | 0.95, 1.14 | 2,339 (43.42%) | 3,048 (56.58%) | 1.19 | 1.05, 1.35 | 1,560 (51.18%) | 1,488 (48.82%) | 1.04 | 0.87, 1.24 |
| Rich | 6,346 (51.48%) | 5,980 (48.52%) | 1.07 | 0.97, 1.18 | 2,180 (36.45%) | 3,800 (63.55%) | 1.28 | 1.12, 1.45 | 1,956 (51.46%) | 1,844 (48.54%) | 1.01 | 0.85, 1.22 |
| Richest | 6,456 (47.45%) | 7,149 (52.55%) | 1.13 | 1.02, 1.24 | 1,837 (25.70%) | 5,312 (74.30%) | 1.37 | 1.20, 1.56 | 2,702 (50.87%) | 2,610 (49.13%) | 1.07 | 0.89, 1.29 |
| **Smoker Status** |  |  |  |  |  |  |  |  |  |  |  |  |
| Has never smoked | 23,260 (48.58%) | 24,620 (51.42%) | Ref. |  | 9,311 (37.82%) | 15,308 (62.18%) | Ref. |  | 7,725 (50.47%) | 7,583 (49.53%) | Ref. |  |
| Former smoker | 2,244 (66.94%) | 1,108 (33.06%) | 0.95 | 0.86, 1.04 | 612 (55.24%) | 496 (44.76%) | 1 | 0.87, 1.13 | 221 (44.51%) | 275 (55.49%) | 0.98 | 0.80, 1.18 |
| Currently or Daily smoker | 3,861 (66.62%) | 1,935 (33.38%) | 0.94 | 0.87, 1.01 | 985 (50.92%) | 950 (49.08%) | 0.92 | 0.84, 1.02 | 467 (49.19%) | 482 (50.81%) | 0.89 | 0.77, 1.04 |
| **Alcohol consumption** |  |  |  |  |  |  |  |  |  |  |  |  |
| Never or did not use in the last 12 months | 8,164 (43.47%) | 10,618 (56.53%) | Ref. |  | 3,615 (34.04%) | 7,004 (65.96%) | Ref. |  | 3,767 (53.79%) | 3,237 (46.21%) | Ref. |  |
| Non-excessive alcohol consumption | 20,236 (55.17%) | 16,441 (44.83%) | 0.94 | 0.90, 0.98 | 7,002 (42.59%) | 9,439 (57.41%) | 0.93 | 0.89, 0.98 | 4,482 (47.49%) | 4,957 (52.51%) | 1.06 | 0.99, 1.14 |
| Excessive alcohol consumption | 964 (61.51%) | 603 (38.49%) | 0.85 | 0.75, 0.96 | 292 (48.43%) | 311 (51.57%) | 0.84 | 0.70, 0.98 | 164 (52.66%) | 147 (47.34%) | 0.93 | 0.71, 1.20 |
| **Fruit and/or vegetable consumption** |  |  |  |  |  |  |  |  |  |  |  |  |
| < 5 servings per day | 26,735 (51.32%) | 25,363 (48.68%) | Ref. |  | 10,030 (39.55%) | 15,333 (60.45%) | Ref. |  | 7,752 (50.56%) | 7,581 (49.44%) | Ref. |  |
| ≥ 5 servings per day | 2,629 (53.35%) | 2,299 (46.65%) | 0.95 | 0.89, 1.02 | 879 (38.22%) | 1,420 (61.78%) | 1.07 | 0.99, 1.16 | 661 (46.55%) | 759 (53.45%) | 1 | 0.88, 1.12 |
| **Nutritional Status** |  |  |  |  |  |  |  |  |  |  |  |  |
| Normal weight | 6,659 (52.40%) | 6,049 (47.60%) | Ref. |  | 2,595 (42.89%) | 3,455 (57.11%) | Ref. |  | 1,661 (48.07%) | 1,794 (51.93%) | Ref. |  |
| Overweight | 12,128 (52.60%) | 10,929 (47.40%) | 0.96 | 0.91, 1.02 | 4,219 (38.60%) | 6,710 (61.40%) | 0.98 | 0.91, 1.05 | 3,329 (49.61%) | 3,381 (50.39%) | 1 | 0.90, 1.12 |
| Obesity | 10,575 (49.76%) | 10,678 (50.24%) | 0.96 | 0.90, 1.03 | 4,091 (38.31%) | 6,587 (61.69%) | 0.96 | 0.88, 1.05 | 3,423 (51.97%) | 3,163 (48.03%) | 0.92 | 0.81, 1.04 |
| **Abdominal Obesity** |  |  |  |  |  |  |  |  |  |  |  |  |
| Normal | 8,427 (61.37%) | 5,304 (38.63%) | Ref. |  | 2,264 (42.69%) | 3,040 (57.31%) | Ref. |  | 1,547 (50.90%) | 1,492 (49.10%) | Ref. |  |
| Obesity | 8,990 (46.22%) | 10,461 (53.78%) | **1.11** | **1.05, 1.17** | 3,750 (35.84%) | 6,711 (64.16%) | **1.11** | **1.04, 1.20** | 3,431 (51.13%) | 3,280 (48.87%) | 0.93 | 0.84, 1.04 |
| **T2DM** |  |  |  |  |  |  |  |  |  |  |  |  |
| No | 27,883 (54.53%) | 23,251 (45.47%) | Ref. |  | 9,783 (42.08%) | 13,468 (57.92%) | Ref. |  | 6,658 (49.44%) | 6,810 (50.56%) | — | — |
| Yes | 1,460 (24.95%) | 4,392 (75.05%) | **1.48** | **1.41, 1.55** | 1,117 (25.44%) | 3,275 (74.56%) | **1.15** | **1.09, 1.22** | 1,751 (53.46%) | 1,524 (46.54%) | 0.93 | 0.86, 1.01 |
| **Race** |  |  |  |  |  |  |  |  |  |  |  |  |
| Other | 10,715 (48.80%) | 11,244 (51.20%) | Ref. |  | 4,720 (41.98%) | 6,524 (58.02%) | Ref. |  | 3,216 (49.30%) | 3,308 (50.70%) | Ref. |  |
| Quechua | 4,690 (53.99%) | 3,997 (46.01%) | 1.09 | 1.00, 1.18 | 1,921 (48.08%) | 2,075 (51.92%) | **0.87** | **0.79, 0.96** | 967 (46.61%) | 1,108 (53.39%) | **1.2** | **1.03, 1.40** |
| Aymara | 433 (56.89%) | 328 (43.11%) | 0.97 | 0.83, 1.14 | 200 (61.03%) | 128 (38.97%) | **0.67** | **0.52, 0.85** | 45 (35.25%) | 83 (64.75%) | **1.25** | **0.89, 1.72** |
| Native or Indigenous Amazonian | 174 (64.04%) | 98 (35.96%) | 0.94 | 0.69, 1.24 | 50 (50.80%) | 48 (49.20%) | 0.93 | 0.59, 1.39 | 28 (59.07%) | 20 (40.93%) | 1.05 | 0.56, 1.79 |
| Negroid/Black | 2,029 (51.98%) | 1,874 (48.02%) | **1.12** | **1.02, 1.23** | 686 (36.60%) | 1,188 (63.40%) | 1.01 | 0.91, 1.13 | 640 (53.85%) | 548 (46.15%) | 1.06 | 0.90, 1.25 |
| White/Caucasian | 1,477 (52.79%) | 1,321 (47.21%) | 1.07 | 0.97, 1.18 | 417 (31.56%) | 904 (68.44%) | 1.06 | 0.94, 1.18 | 530 (58.57%) | 375 (41.43%) | 0.94 | 0.79, 1.12 |
| Mestizo | 9,846 (52.81%) | 8,800 (47.19%) | **1.11** | **1.03, 1.19** | 2,914 (33.11%) | 5,886 (66.89%) | 0.95 | 0.87, 1.04 | 2,986 (50.74%) | 2,900 (49.26%) | 1.1 | 0.96, 1.26 |
| **Altitude** |  |  |  |  |  |  |  |  |  |  |  |  |
| 0 to 499 | 20,092 (51.76%) | 18,722 (48.24%) | Ref. |  | 6,268 (33.48%) | 12,454 (66.52%) | Ref. |  | 6,436 (51.67%) | 6,019 (48.33%) | Ref. |  |
| 500 to 1499 | 2,014 (50.82%) | 1,949 (49.18%) | 0.93 | 0.86, 1.01 | 889 (45.63%) | 1,060 (54.37%) | 0.91 | 0.82, 1.01 | 476 (44.91%) | 584 (55.09%) | 1.12 | 0.97, 1.29 |
| 1500 to 2999 | 3,260 (50.30%) | 3,221 (49.70%) | **0.81** | **0.70, 0.94** | 1,474 (45.78%) | 1,746 (54.22%) | 0.83 | 0.67, 1.01 | 910 (52.10%) | 837 (47.90%) | 0.85 | 0.63, 1.14 |
| 3000 or more | 3,998 (51.46%) | 3,771 (48.54%) | 0.87 | 0.74, 1.03 | 2,277 (60.39%) | 1,493 (39.61%) | **0.67** | **0.53, 0.85** | 592 (39.62%) | 902 (60.38%) | 1.1 | 0.78, 1.54 |

Each factor has been independently adjusted for Sex, Group Age, Educational Level, Civil Status, Natural Region, Area Of Residence, Wealth Index, Smoker Status, Alcohol Consumption, Fruits and/or Vegetable Consumption, Nutricional Status, Abdominal Obesity, T2DM, Race, and Altitude
aPR: Prevalence ratio. CI 95%: Confidence interval at 95%
Source: self-made

### Prevalence of HTN, Prehypertension, and HBP

The global prevalence of HTN was 20.76%. Analyzing the annual trend, it was observed that the prevalence of HTN was 21.97% in 2014, decreasing the following year but then gradually increasing to 23.38% in 2022.

Regarding prehypertension, breaking down the data annually, the prevalence of prehypertension was 33.13% in 2016, experiencing fluctuations over the years, with its most significant increase in 2021 and decreasing to 32.69% in 2022. On the other hand, the provided data indicate that the prevalence of HBP was 16.52% in 2016 and has experienced a gradual increase over the years, reaching 22.26% in 2022.

### Prevalence of Awareness, Medicated, and Control HTN

The global Awareness prevalence of HTN was 48.51%. Observing the annual trend, the prevalence of Awareness HTN was 50.42% in 2016 and has shown some fluctuations over the years, being 49.00% in 2022 (Figure 1).

**Figure 1.**
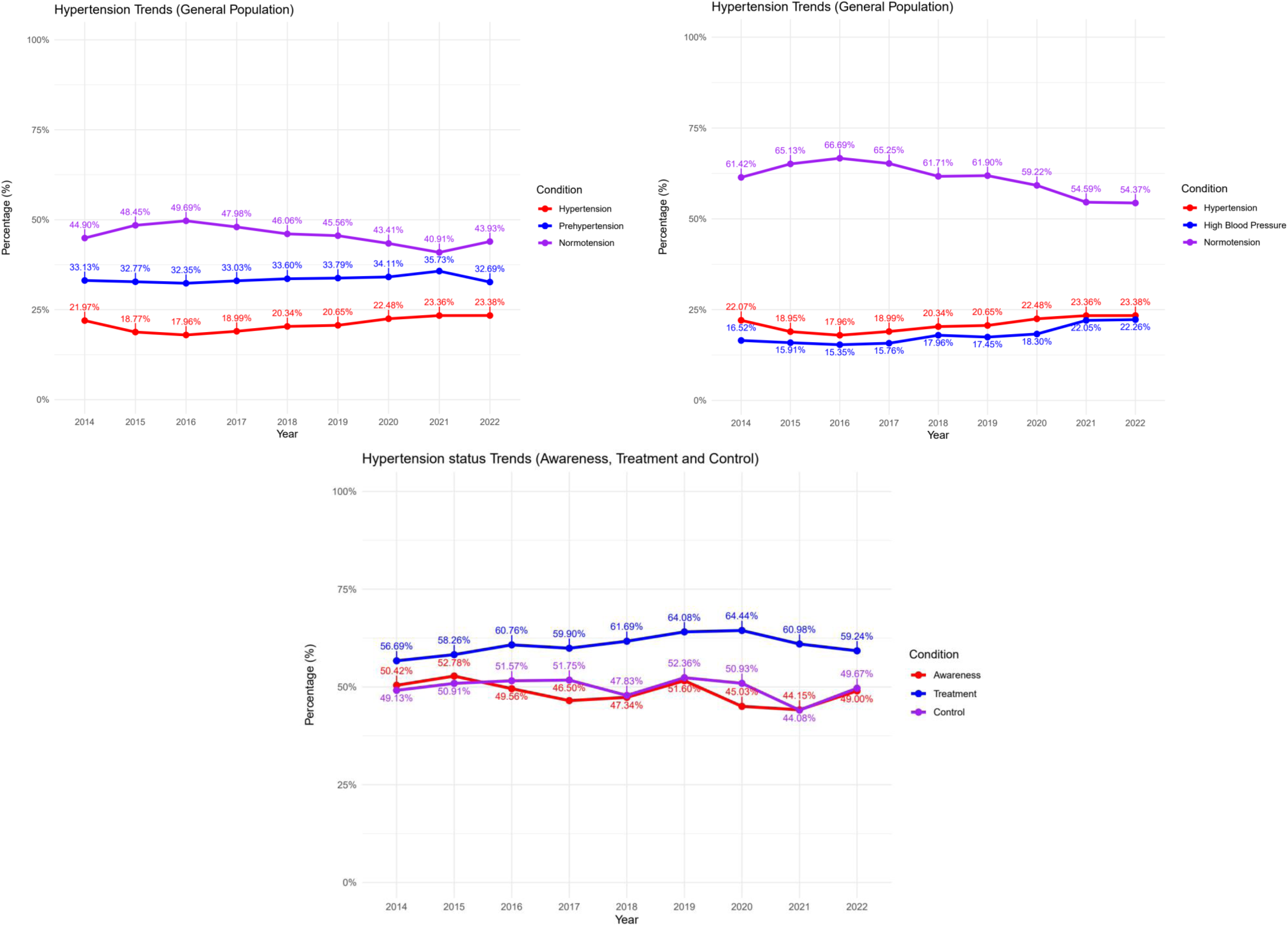
Trend graph of types of arterial hypertension (normotension, prehypertension, HBP and HTN) and HTN states (Awareness, Treatment and Control)

In the case of TreatmentHTN, its total prevalence was 60.56%. Breaking down the data by year, it was observed that the prevalence of TreatmentHTN has varied over time, being 56.69% in 2016 and 59.24% in 2022 (Figure 1).

Finally, 49.78% of these individuals indicated having their HTN under control. Analyzing the data by year, it was observed that the prevalence of Control HTN has fluctuated over time, ranging between 49.13% in 2021 and 49.67% in 2019 (Figure 1).

### Factors Associated with HTN

Our analysis of factors associated with an elevated HTN occurrence revealed that males exhibited a higher prevalence in relation to females. Those of more advanced age, especially those over 35, showed a higher occurrence, with the risk growing substantially in individuals surpassing seventy years. Educational levels uncovered a pattern whereby persons possessing greater educational attainment showed a decreased frequency of HTN as opposed to individuals with little or no official instruction. Married or partnered individuals had a lower prevalence of HTN than their single counterparts. Geographically, residents of other coastal areas and the highlands exhibited a lower prevalence of HTN than those in Metropolitan Lima, whereas the jungle regions showed a distinctly lower prevalence. Moderation in alcohol consumption was linked to a lower prevalence of HTN compared to higher consumption levels. Those who were overweight or obese exhibited a more significant occurrence of high blood pressure contrasted with individuals of average size, whereby excess abdominal fat served to amplify this occurrence to an even higher degree. The presence of T2DM was associated with a higher prevalence of HTN. While ethnicity proved influential, those identifying as Aymara exhibited a decreased occurrence of other ethnic affiliations in the study. Furthermore, residing at moderate altitudes from 500 to 1499 meters appeared to correlate with reduced high blood pressure in lower elevations.

### Factors Associated with Prehypertension and HBP

Upon investigating prehypertension, it was discerned that the prevalence among males exceeded that found for females. While those between 36-59 years of age and 60-69 years exhibited a heightened prevalence compared to the younger group of 18-35, groups of 70 or more displayed a somewhat reduced prevalence. Other than Metropolitan Lima and the highlands, residents of the coastal areas showed a lower prevalence, while the jungle region revealed a higher prevalence. Those in relationships had a lower occurrence of prehypertension when compared to singles, according to how being coupled correlated with a decreased prevalence of HTN versus being uncoupled. Those with primary, secondary, or higher educational attainment demonstrated a decreased occurrence compared to individuals without schooling. Overweight and obese individuals, as well as those with T2DM, exhibited a higher prevalence, according to the data. Those of Aymara ethnicity and residents at altitudes between 1500 and 2499 meters showed a higher prevalence than other groups.

When focusing on HBP, men again showed a significantly higher prevalence than women. The prevalence among those aged 36-59, 60-69, and 70 years or older proved to be higher compared to individuals in the 18-35 age bracket. Living outside of Metropolitan Lima, specifically in other coastal areas, the highlands, and the jungle, was associated with a lower prevalence. Being in a relationship, living in rural areas, and belonging to wealthy groups, poor, middle, rich, and most affluent, indicated a lower prevalence than the poorest group. Those who consumed alcohol excessively or had a higher intake of fruits and vegetables were more likely to have a higher prevalence, as was noted. Those who were overweight, obese, or had T2DM, especially individuals of Caucasian or Mestizo descent, exhibited a notably higher occurrence than other groups, according to the findings. Residing at altitudes of 1500 to 2999 meters and 3000 meters or more was associated with an increased prevalence of HBP.

### Factors Associated with Awareness HTN

When considering factors related to Awareness of HTN, it was observed that regarding sex, males exhibited higher hypertension levels in contrast with females. As age groups progressed to more advanced stages, the probability of developing this condition increased in correlation to aging. Educational levels showed a trend whereby higher degrees corresponded to a lower likelihood of HTN occurrence. The same was observed in individuals who were in a relationship. Regarding region, people residing in jungle areas had a lower prevalence. However, research also indicated that regular but moderate alcohol intake was correlated with an elevated likelihood of contracting the condition. Those with higher weights tended to have more excellent rates of hypertension when considering nutritional conditions, as overweight and obese individuals exhibited more prevalent instances of elevated blood pressure. The relationship between abdominal obesity and T2DM similarly held constant. Additionally, the Aymara ethnicity showed a lower prevalence compared to other groups. Living at higher altitudes between 500 and 1499 meters correlates with a lower prevalence of HTN.

### Factors Associated with Treatment HTN

In the context of the treatment of HTN, Age is a significant factor. As one advances in age, especially those beyond their mid-thirties, the probability increases of requiring pharmaceutical intervention to regulate blood pressure, with this phenomenon being most pronounced among those who have surpassed their seventh decade. Being in a relationship seems to have a protective effect associated with lower rates of Treatment HTN. Your economic status also comes into play. Interestingly, those from the poorer to the most affluent categories report higher hypertension rates than the poorest group. Having abdominal obesity or Type 2 Diabetes also makes it more likely that you’ll be on medication for hypertension. And there’s a fascinating link with ethnicity – people from Quechua or Aymara backgrounds tend to have lower rates. Also, living higher up, between 1500 to 2999 meters or even above 3000 meters, reduces the need for HTN medication.

### Factors Associated with Control HTN

Regarding Control HTN, men showed a lower prevalence compared to women. As age increased, particularly in age groups over 36 years, a lower prevalence was observed, with this trend being more pronounced in those over 70 years. Geographical location was influential, with residents living on the coast outside of Metropolitan Lima and in the jungle regions showing a higher prevalence. Ethnicity, particularly Quechua ethnicity, was associated with a higher prevalence of Control HTN.

## Discussion

### Prevalence of Hypertension in Peru

The prevalence of HTN in Peru has exhibited an upward trend between 2014 and 2022, peaking notably around 2022. This increase is concerning and reflects a growing trend of HTN, not only nationally but also in the Latin American and global context. The WHO reports that the worldwide occurrence of high blood pressure has been increasing, affecting approximately one-third of all adults, with a higher frequency found in poorer developing nations ^(13)^. Recent studies have comparably shown that the prevalence of high blood pressure in Latin American adults fluctuates within a range of 20% to 40%, with numerous nations exhibiting figures that match or perhaps surpass what has been seen regarding such matters within Peru precisely ^(19)^.

A multitude of influences have led to the rising occurrence of HTN in Peru, including evolving lifestyles, a growing prevalence of obesity and diabetes as potential risk factors, and enhanced disease diagnosis. Furthermore, the aging population’s potential contribution to such a rise cannot be discounted ^(6)^. These trends emphasize the necessity of implementing astute public health tactics for both preempting and regulating high blood pressure in Peru, taking into account its consequences on infirmity and death associated with cardiovascular diseases.

### Prevalence of Prehypertension and High Blood Pressure (HBP) in Peru

Although the latest medical guidelines have replaced prehypertension with HBP, it is crucial to consider both terms in the study’s context. For quite some time now, prehypertension has endured widespread application and utilization, maintaining significant applicability toward comprehending the progressive development and handling of conditions characterized by elevated arterial blood pressure ^(14)^. In Peru, the prevalence of prehypertension, observed over the years, reflects a significant public health concern. This phenomenon, which aligns with global and Latin American trends where research has found that between 30% and 50% of adults in various areas have been identified as prehypertensive, reflects reports that an equivalent percentage falls into that category ^(20)^.

Recognizing prehypertension as a risk state for developing HTN is vital for implementing preventive strategies. Despite changes in terminology, focusing on prehypertension underscores the importance of early detection and intervention to prevent progression to HTN ^(21–23)^. On the other hand, ABP offers a more dynamic assessment of blood pressure, allowing for early identification of those at risk ^(24,25)^.

Including prehypertension and HBP in studies and public health, strategies are fundamental, as they reflect the changing nature of understanding and managing HTN. Analyzing these categories is crucial for effective and early intervention in the Peruvian population, thus mitigating the risk of complications associated with hypertension.

### Factors Associated with HTN

The factors associated with HTN in Peru, identified in this study, offer an intriguing panorama when contrasted with trends observed in Latin America, the United States, and globally. In Peru, factors such as gender, advanced age, low educational level, marital status, geographic region, and the presence of comorbidities like obesity and T2DM are significantly associated with the prevalence of HTN. These findings align with patterns observed in the other areas, albeit with some notable variations.

Globally and in Latin America, studies have demonstrated that HTN is strongly associated with similar risk factors, including the aging population, rising obesity, and sedentary lifestyles ^(13)^. In the United States, HTN is also related to age, gender, race/ethnicity, and socioeconomic factors. However, prevalence and associated factors may vary due to differences in lifestyle, diet, and access to healthcare ^(26)^.

A fascinating comparison regards metabolic criteria, such as obesity and diabetes. Both in Peru and globally, obesity has been established as a significant risk factor for HTN ^(27,28)^. In Latin America, the increasing prevalence of obesity has paralleled the rise in HTN, suggesting an interrelation between these conditions ^(19,29)^. In the United States, this relationship is also evident. However, the impact of additional factors like race/ethnicity and socioeconomic disparities plays a crucial role in the prevalence and management of HTN^(30)^.

It is crucial to note that while there are similarities in HTN risk factors between Peru, other Latin American regions, the United States, and globally, intervention and prevention strategies must be adapted to the specific context of each region. This involves considering cultural, socioeconomic, and health service access differences in planning and executing public health programs to combat hypertension.

### Factors Associated with Prehypertension and HBP

The factors associated with prehypertension and HBP in our study show similarities and differences with those related to HTN. In the context of the former two, factors such as gender, age, lifestyle, and comorbidities emerged as influential. These findings align with international studies, where risk factors for prehypertension and HBP often overlap with those for HTN ^(31,32)^.

In terms of similarities, both for prehypertension and HTN, factors like advanced age, overweight/obesity, and T2DM are consistently reported as high risk. However, there is a notable difference in the influence of gender and ethnicity, which may vary between prehypertension and HTN, as indicated by studies conducted in different regions, including Latin America and the United States ^(33–35)^.

The progressive nature of the disease can explain the relationship between factors associated with prehypertension/ HBP and HTN. Prehypertension and HBP are often considered precursor stages of HTN, where the same risk factors may contribute to developing a more severe condition. However, the absence of a direct correlation in some cases may be due to variations in genetics, environment, and lifestyle, which influence the early and late stages of hypertension differently ^(35,36)^.

It is essential to highlight that while some factors are shared between prehypertension/ HBP and HTN, how these factors interact and contribute to disease progression may vary. For example, the impact of diet and exercise might be more pronounced in the early stages (prehypertension/HBP) compared to later stages of HTN, where pharmacological treatment and comorbidity management become more relevant ^(37,38)^.

### Prevalence and Trends of Known, Medicated, and Controlled HTN

The prevalence and trends of known, medicated, and controlled HTN in Peru reveal essential aspects of this condition’s management. Our study observed that, although a significant proportion of individuals with HTN were aware of their condition, not all received pharmacological treatment, and only a minor percentage achieved effective blood pressure control. This situation reflects similar global challenges in HTN management.

While global reports have revealed that merely a minor fraction of people experiencing HTN are aware of their condition, seek medical attention, and effectively control their blood pressure levels, improved education and preventative care appear vital to alleviating this widespread public health concern. According to a WHO study, fewer than half of individuals with HTN in lower and middle-income nations are aware of their condition, with an even smaller portion obtaining treatment and achieving control as indicated ^(4)^. Likewise, in Latin America, a sizable share of those afflicted by high blood pressure have gone undiagnosed, remain without treatment, or are ineffectively managing the condition ^(19)^.

Compared to countries like the United States, where awareness, treatment, and control of HTN are generally higher due to better healthcare access and awareness campaigns, Peru faces particular challenges regarding health resources, access to medications, and health education ^(30)^. This underscores the necessity of implementing nuanced public health strategies in Peru that center on detecting and treating HTN and educating and aiding patients to enhance long-term commitment and management through continual support.

The trend observed in Peru, where the prevalence of known HTN has shown fluctuations, and treatment and control remain suboptimal, suggests the need to strengthen health systems and public health policies. These measures should focus on improving access to diagnostics and treatments and educating the population about the importance of HTN management and the risks associated with lack of control.

### Factors Associated with Known, Medicated, and Controlled HTN

The factors associated with known, medicated, and controlled HTN in Peru present similarities and differences. In our study, aspects such as age, gender, socioeconomic level, access to medical care, and comorbidities played a significant role in these HTN categories. Some exciting correlations emerge when comparing these findings with studies conducted in Latin America, the United States, and globally.

Typical factors for known HTN include advanced age and access to medical care. Studies in Latin America and the United States have shown that better access to healthcare services translates into greater awareness of the condition ^(19,39)^. Additionally, in Peru, as in other contexts, men tend to be less aware of their hypertension condition compared to women.

Regarding medicated HTN, age and comorbidities, such as diabetes and obesity, are relevant factors both in Peru and globally. Studies indicate that patients with comorbidities are more likely to receive treatment for HTN, a pattern also observed in the United States and other regions ^(5,40)^.

For controlled HTN, treatment adherence is crucial. In this aspect, factors like education level and social support play an essential role, similar to those observed in other countries ^(41–43)^. The difficulty in controlling HTN despite treatment may be related to genetic factors, lifestyle, and the presence of concomitant diseases.

It is important to note that while there are similarities in the factors associated with known, medicated, and controlled HTN between Peru and other regions, intervention strategies must be adapted to the specific context of each country or region. This includes considering differences in the healthcare system, access to medications, and cultural factors that may influence the perception and management of hypertension.

### Importance of the Results for Public Health

The findings of this study on HTN in Peru have significant implications for the country’s public health policies. The growing prevalence of HTN, along with observed trends in prehypertension, HBP, and known, medicated, and controlled HTN, highlights the urgent need for comprehensive health strategies. These strategies should focus on detection and medical treatment, health education, promoting healthy lifestyles, and access to quality healthcare services. Implementing public health programs that address these factors can significantly reduce the burden of cardiovascular diseases and other complications associated with HTN, thus improving the quality of life of the Peruvian population ^(44)^.

The findings underscore the importance of effective prevention and control of HTN. Early identification of individuals at risk, such as those with prehypertension or HBP, is crucial for implementing preventive interventions and preventing progression to HTN. Additionally, improving awareness and management of known, medicated, and controlled HTN can significantly impact reducing morbidity and mortality related to this condition. These actions require a collaborative approach involving healthcare professionals, health policymakers, and the community ^(45)^.

Globally, the results of this study can serve as a reference for other countries, especially those in development with similar challenges in HTN management. Understanding the associated factors and trends in HTN can guide the formulation of public health policies and programs tailored to the specific needs of each region. Additionally, the comprehensive approach adopted in Peru could be a model for other countries seeking to improve HTN control and its associated complications ^(46)^.

## Limitations of the Study

While the cross-sectional design of this study precludes determining causal connections between identified risk aspects and hypertension, its transversal nature significantly restricts establishing relationships between the factors seen and high blood pressure over time. The cross-sectional design only allows for observing associations at a specific time, meaning trends or changes in individuals’ health status over time cannot be tracked. This limits the study’s ability to identify the temporal sequence of exposure to risk factors and the development of HTN.

A further pertinent constraint pertains to the dependence on self-reported information for specific factors, such as conformity to remedy and lifestyle habits. While self-reported information risks inaccuracies stemming from recollection flaws or tendencies to portray oneself positively, such factors could compromise the outcomes’ veracity.

While the study examines a broad cross-section of Peruvian residents, its findings may have limited external validity and not fully translate to other settings or groups due to alternative regional influences. Variances regarding culture, socioeconomic class, medical care availability, and living habits can impact hypertension’s frequency and treatment in a region, confining the power to apply the results elsewhere internationally or locally broadly.

While the research may have left out some critical variables impacting high blood pressure, such as genetic, ecological, or psychosocial aspects, a more comprehensive assessment incorporating additional pertinent influences could potentially furnish valuable insights into better understanding and addressing this significant health issue. Omitting these variables could affect understanding of the factors associated with HTN and management.

Finally, blood pressure measurement can be affected by variations in measurement technique, equipment used, and environmental conditions during the measurements. Although standardized protocols were followed, this type of study’s variability in blood pressure measurement is an inherent limitation.

## Conclusions

The investigation into HTN in Peru revealed an expanding occurrence of HTN, prehypertension, and HBP, emphasizing the necessity for early identification, suitable treatment, and effective management of HTN. A significant gap is observed between the condition’s awareness and effective management, highlighting the urgent need for public health strategies that address both the prevention and treatment of HTN in Peru.

Strengthening health prevention and education programs, focusing on healthy lifestyles to mitigate HTN risk factors, is essential. Improvements in access to early detection and diagnosis and strategies to increase treatment adherence are crucial. Additionally, formulating inclusive policies that consider the social determinants of health and promoting ongoing research to guide and optimize these interventions is necessary.

The findings of this study have global implications, providing a framework for other countries with similar challenges in HTN management. Longitudinal studies are recommended for a deeper understanding of HTN and assessing the long-term impact of various management strategies. Multisectoral collaboration and an integrated approach are essential for effectively addressing HTN in Peru and worldwide.

## Acknowledgments

A special thanks to the members of Instituto de Investigación de Enfermedades Tropicales, Universidad Nacional Toribio Rodríguez de Mendoza de Amazonas (UNTRM), Amazonas, Perú who provided valuable comments during the preparation of this study.

## Informed consent

It was not necessary to obtain informed consent in this Study

## Data availability

The data supporting the findings of this study can be accessed by the original research paper at the follow link: https://proyectos.inei.gob.pe/microdatos/

## Author contributions

**Víctor Juan Vera-Ponce:** Methodology, Data Analysis, Writing – Review & Editing.

**Fiorella E. Zuzunaga-Montoya:** Conceptualization, Data Analysis, Writing – Review & Editing.

**Joan A. Loayza-Castro:** Data Analysis, Methodology, Validation, Writing – Original Draft.

**Luisa Erika Milagros Vásquez Romero:** Data Analysis, Methodology, Validation, Writing – Original Draft.

**Cori Raquel Iturregui Paucar:** Validation, Visualization, Writing – Review & Editing.

**Mario J. Valladares-Garrido:** Project Administration, Supervision, Methodology, Writing – Review & Editing.

**Enrique Vigil-Ventura:** Supervision, Methodology, Funding Acquisition, Writing – Review & Editing.

